# From community as data providers to community as data users: developing a community-led research platform using program data in HIV/STI Program Science in Kenya

**DOI:** 10.1101/2024.12.03.24318454

**Authors:** Nancy Tahmo, Anthony Noah, Byron Odhiambo, Charles Kyalo, Elly Ondiek, Fortune Ligare, Gilbert Asuri, Jedidah Wanjiku, John Alex Njenga, John Maina, Kennedy Mwendwa, Kennedy Olango, Kennedy Ouma, Loice Nekesa, Pascal Macharia, Silvano Tabbu, Kristy CY Yiu, Robert Lorway, Parinita Bhattacharjee, Huiting Ma, Lisa Lazarus, Sharmistha Mishra, Jeffrey Walimbwa, Health Research Intervention Kuthamini Afya YeCommunity-Led Research Initiative

**Author notes:** Corresponding author: (JW); (NT). Co-senior authors (Sharmistha Mishra, Jeffrey Walimbwa).

## Abstract

**Background:** Community-based organizations (CBOs) are critical in providing trusted and targeted HIV/STI services to gay, bisexual, and other men who have sex with men (GBMSM). Despite significant strides in CBOs’ involvement in HIV/STI research, there remain gaps in meaningful engagement, especially in quantitative research. This paper explores the development of HEKA, a community-led research platform where community-based organizations build capacity and leverage routinely collected program data to design research that aims to improve HIV/STI programs. We share a collective reflection on the lessons learned in the process, the challenges that emerged, and recommendations for facilitating community-based program science.

**Methodology:** Through a collaborative process, seven CBOs serving GBMSM in Kenya created the HEKA Research Initiative and designed a framework of collaboration, through which we assessed the technical gaps in quantitative research among staff, applied for funding, co-designed capacity-building workshops with academic partners, and developed a research agenda. We established a monthly meeting frequency and through collective reflection, documented the lessons and challenges in the process.

**Outcomes:** With our successful grant, we organized an in-person workshop on quantitative research methods and R programming. The team identified research questions and completed data cleaning/harmonization of program data. HEKA was successful because we emphasized a co-leadership framework (research direction evolved through shared/delegated leadership), and peer-to-peer mentorship. Major challenges included: obtaining sustained funding for engagement; ensuring the learning pace allows all individuals to be on the same page; confronting the socio-political climate; long commutes between counties for in-person meetings; and the limitation in using Excel files as primary tools for data capture.

**Conclusions:** HEKA demonstrates the potential for community-based and led research in the HIV/STI field. The model we present can serve as a blueprint for other community-based organizations aiming to lead collaborative or independent research and build capacity.

## Introduction

Since the early days of the HIV epidemic, communities have demanded to play an active role in HIV research(1). HIV activists have challenged the portrayal of people living with HIV as “victims”, shifting the narrative to self-empowerment and community-led HIV research(2). Community involvement has advanced HIV research, with prominent examples like the Community Advisory Board of the Advancing Clinical Therapeutics Globally for HIV/AIDS and Other Infections (formerly AIDS Clinical Trials Group)(3). In 2007, the Joint United Nations Programme on HIV/AIDS developed the Good Participatory Practice framework rooted in community-based participatory research and intended to foster power-balanced and transparent community involvement within HIV biomedical prevention trials(4). There have been some strides in collaboration within clinical research and the social sciences, such as the recently completed polling booth survey where community members acted as data collectors(5). However, there remains gaps in meaningful community engagement in many quantitative fields like mathematical modeling of HIV/STI, with concerns surrounding community involvement as a checkbox(6).

A recent review revealed gaps in meaningful community and academic collaboration(7). This is particularly so around alignment between academics’ research goals and the priorities of community programs (“democratic deficit”), compensation for communities, and evaluation of the effect of such collaborations on program delivery(7). Many reports on community engagement provide a narrative of experiences and lessons learned (usually a researcher-curated viewpoint), with fewer reports appraising their effectiveness in adding value to the delivery of community programs and health outcomes (the community viewpoint)(4,7). Also, more often, the paternalistic approach to research is such that a community identifies local issues and academic researchers attempt to resolve them through research with communities (in which case, technical skills are often attributed to the academic researchers). The problem with this approach is that the community organizations fall behind in research capacity to conduct assessments using program data, adapt program delivery, and independently secure competitive funding(8). The traditional solution has been the use of short one-time training sessions with community program staff (mostly geared at research partnerships with academics)(8). There have been attempts at developing community-led research networks in Kenya focused on quantitative research, such as G10 through the International AIDS Vaccine Initiative (IAVI). However, G10’s capacity has mostly been as an advisory board for quantitative research(9).

Against this backdrop, the program science framework was developed to offer an innovative and adaptive solution. Program science systematically encourages a bi-directional approach where program implementers inform research, and research, in turn, informs program implementation and policy in an iterative process(10). This framework has the potential to unify community-academic researcher collaborations to inform the three pillars of program decisions: strategic planning (e.g., understanding socio-epidemiological drivers of transmission in epidemics of HIV and other sexually transmitted infections [STI]); program implementation (e.g., prioritizing interventions and reaching effective coverage); and program evaluation (e.g., monitoring for impact on health and wellbeing). As such, program science also uniquely positions community-based organizations to meet the hallmarks of community-led monitoring, an evolving area of HIV/STI prevention with evidenced health outcome improvement(11). Community-led monitoring is an iterative process where communities actively participate in co-designing how data is collected, analyzed, and leveraged to improve the delivery of programs and improve health outcomes(12).

As in many countries in Southern and Eastern Africa, there is a high prevalence of HIV/STI in Kenya(13). In this context, operationalizing HIV/STI program science and community-participatory research in HIV/STI prevention and treatment in Kenya has largely been in social science and qualitative research(14,15). These include but are not limited to, building capacity in qualitative data collection such as project ethnography, developing, translating, and conducting interviews, and thematic analyses. When quantitative methods are used in program science, such as cross-sectional surveys or analyses of programmatic cohort data, community partners have been engaged through consultation to develop routine data collection tools and/or to design interventions(10). Often this takes the form of community advisory boards or community expert advisory groups and data collectors.

Yet in Kenya, as in many other settings, community-based organizations also implement programs providing front-line services for HIV/STI prevention and treatment. Box 1 summarizes the types of services provided by our community-based organizations. These programs also collect routine program data, guided by standardized tools developed by the Government of Kenya National AIDS and STI Control Programme. The program data comprise person-level indicators, with baseline registration information and longitudinal information on each program-client encounter (in clinic or at outreach). At each encounter, services may include HIV testing, STI testing or syndromic assessment, screening for experiences of violence, and risk reduction counseling. Data generated through these services are collected using tools categorized into Community Outreach, Clinical, and Programme-level tools, as shown in Table 1 (Figure 1 details the data flow from encounters with service users to governance and data use). To date, program data have been collected in Excel spreadsheets and submitted to funders and to the National AIDS and STI Control Programme for central data reporting as part of the national monitoring and evaluation system for HIV programmes in Kenya. Programs are starting the transition to electronic medical records (EMR), though some are advanced in the transition and some others beginning the process.

**Table 1.** List of tools used for data collection and handling authorities.

| Category | Name of tool | Handling responsibility |
| --- | --- | --- |
| **Community outreach tools** | Contact form | Peer educator |
|  | Peer educator outreach calendar | Peer educator |
|  | Outreach worker's summary sheet | Outreach worker |
|  | Hotspot listing tool (key populations) | Outreach worker |
|  | Peer educator follow-up tracking form | Peer educators |
|  | Peer LTFU (lost to follow-up) register (key populations) | Peer educator |
|  | Case management forms | Case navigator |
|  | Referral register for social-protection services | Outreach supervisor |
| **Clinical tools** | Clinic enrolment form | Health care worker |
|  | Clinic visit form | Health care worker |
|  | Sexually transmitted infection (STI) treatment data collection form | Health care worker |
|  | Alcohol abuse screening tool | Health care worker |
|  | Patient Health Questionnaire-9 (PHQ-9) for depression screening | Health care worker |
|  | HTS lab & linkage register | Health care worker |
|  | HIV self-testing tracking log | Health care worker |
|  | Partner notification tracking log | Health care worker |
|  | People living with HIV tracker | Health care worker |
|  | Link facility tracking form | Health care worker |
|  | Post-exposure prophylaxis register | Health care worker |
|  | Prep register | Health care worker |
|  | Cervical cancer screening register | Health care worker |
|  | Sex and gender-based violence register | Health care worker |
|  | Treatment preparation register | Health care worker |
|  | Clinical encounter green card | Health care worker |
|  | Art and cohort register | Health care worker |
|  | HIV daily activity register | Health care worker |
|  | Tuberculosis register | Health care worker |
|  | STI treatment tracker-electronic | Health care worker |
|  | Isoniazid Preventive Therapy register | Health care worker |
| **Programme-level tools** | Violence reporting form | Programme officer/advocacy officer/community paralegals |
|  | Training summary tool | Training officer |
|  | Key population master register | Monitoring and evaluation officer |
|  | Drop-in-centre register | Receptionist |
|  | Key population cohort register | Monitoring and evaluation officer |
| **Note.** Find the definitions of each data tool and respective variables here: Key populations programme data collection tools revised reference manual – 2019<br>https://cquin.icap.columbia.edu/wp-content/uploads/2021/08/KP-Tools-Narrative_FINAL.pdf |  |  |

**Figure 1.**
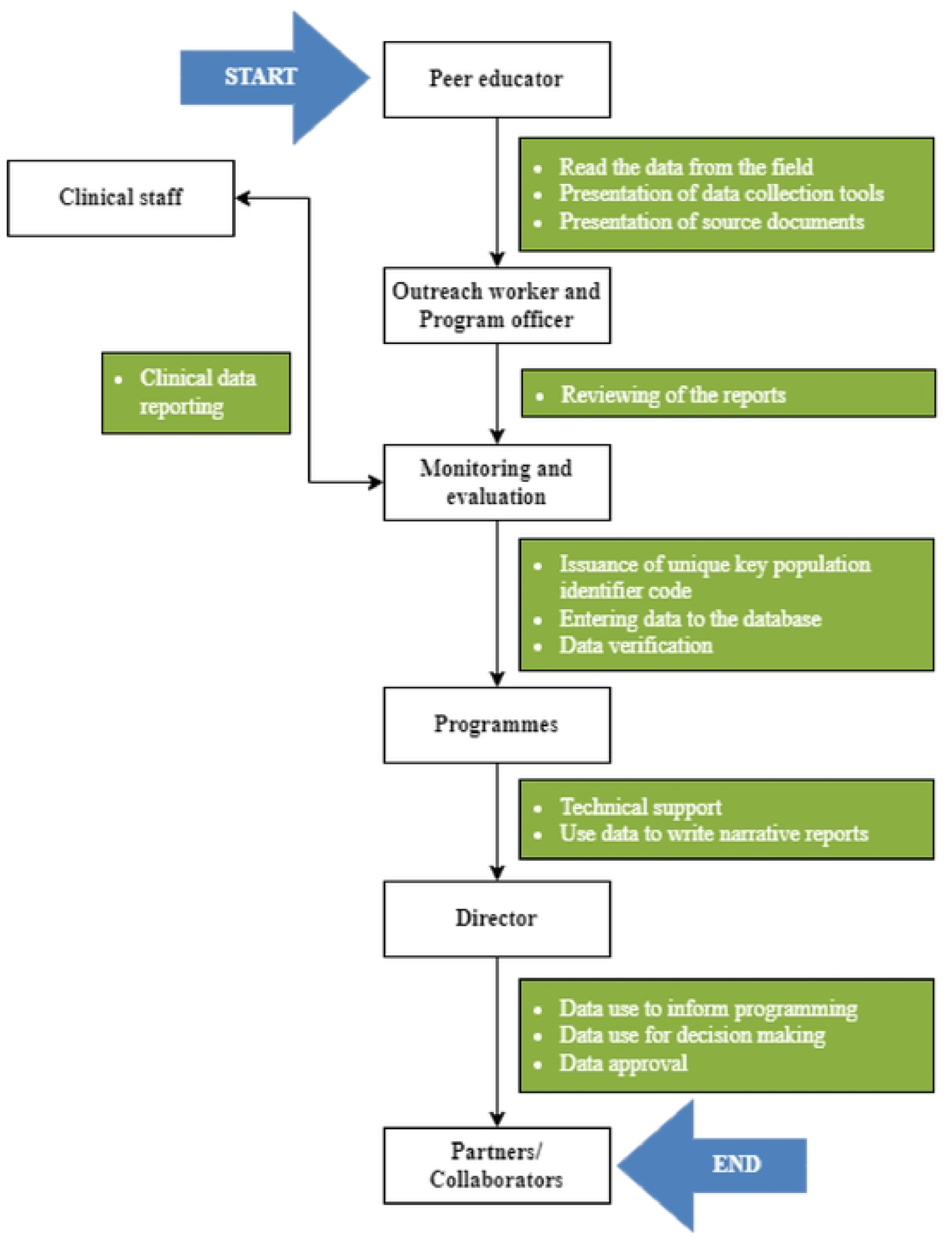
Data flow from service user encounter to governance and data use.

In 2022, a group of seven community-based organizations serving gay, bisexual, and other men who have sex with men (GBMSM) in Kenya came together to form a community-led research initiative to lead quantitative research for community-led program science, using their own program data. This article details the process undertaken by community members in developing HEKA: <u>H</u>ealth R<u>e</u>search Intervention <u>K</u>uthamini <u>A</u>fya Yetu). We share a collective reflection on the lessons learned in the process from the community perspective. Particularly, building community-academic trust; logistics and practical considerations; assessment of the influence of the collaborative process on program delivery; and based on facilitators or barriers that emerged, share pathways for community-led quantitative HIV/STIs research. The initiative paves the way for leveraging routinely collected programmatic data, which aligns with the WHO’s Consolidated guidelines on person-centered HIV strategic information. The WHO highlighted for the first time in 2022, the impact of using programmatic data, as it is reliable in tracking HIV indicators (prevention, testing, and treatment), enhancing timely decision-making, and linkage to STI services(16).

## Methodology

### Site setting

GBMSM in Kenya experience disproportionately high rates of HIV and STI(17–19). The existence of regressive laws and resurgence in public discourse that reinforce discrimination against same-sex sexual practices in Sub-Saharan Africa, and in Kenya particularly, amplifies disparities in health and wellbeing. In Kenya, same-sex sexual practices can result in prison time of up to 14 years(20), compounded by widespread socio-cultural beliefs that provide room for discrimination and violence against GBMSM and sexual minorities. GBMSM report experiencing stigma and discrimination at interpersonal and institutional levels, such as blackmail, constrained healthcare service access, and experiences of violence(21,22). Qualitative studies show that GBMSM in Kenya feel more comfortable utilizing community-based services, and community-based organizations have been associated with effective/targeted approaches to addressing the HIV/STI epidemic(23,24). The HEKA Research Initiative brings together seven community-based organizations that provide HIV/STI prevention, testing, and treatment services to GBMSM in Kenya. Together, we serve six counties: Mombasa, Kilifi, Kisumu, Nakuru, Nairobi, and Uasin Gishu, representing a mix of moderate to high-priority counties(18,25). We came together to develop a collective research platform, utilizing program data to inform our local programs.

### Guiding principles and authors’ positionality

The HEKA research initiative was founded in 2018 under the principles of GIPA/MIPA-Greater/Meaningful Involvement of People Living with HIV/AIDS(26,27) and Community-based Program Science – program coverage framework for HIV/STI research(10,15,28). GIPA/MIPA highlights the importance of the meaningful involvement of people living with or affected by HIV/AIDS in the HIV epidemic response. Complementary to that, community-based program science in HIV/STI coverage emphasizes community collaboration in the iterative use of routine program data to monitor and tailor program delivery.

The HEKA partnership included seven community-based organizations (CBOs): ISHTAR (Nairobi County), HOYMAS (Nairobi), HAPA KENYA (Mombasa), AMKENI-Malindi (Kilifi), KYDESA (Nakuru), Q INITIATIVE (Uasin Gishu and Trans-Nzoia), and MAAYGO (Kisumu). Each CBO has a drop-in center and facilities that provide HIV/STI prevention and care, and services like peer education, counseling, and socio-economic support. The organizations were identified through the GBMSM HIV prevention network of organizations implementing HIV programming in Kenya (GHPN-Ke). Twenty-one community program managers and monitoring and evaluation staff from across the organizations constituted the team. The program managers and monitoring and evaluation staff were included because they are directly involved in coordination and data management in their organizations. Through our shared experiences as program staff— some living with HIV, identifying as sexual/gender minorities, or closely connected to these communities—we have progressively mobilized around program-to-program support. In 2019, the HEKA collective reached out to the Mishra Lab through SM for technical support as an academic collaborator with formal training in epidemiology and mathematical modeling.

SM is a clinician scientist working in the field of mathematical modeling of infectious disease transmission, specifically focusing on HIV and other sexually transmitted diseases. LL, PB, and RL are social scientists working on community-based research in the HIV/STI field, including in Kenya and with some of the HEKA organizational partners. NT is an epidemiologist who has supported HIV/STI programming in a public health department and is serving as the rapporteur for this paper. NT, KCYY, HM, RL, PB, LL, and SM have provided ongoing mentorship and support through research and facilitating capacity-building for our community researchers.

### Ethics statement

We have ethics approval through AMREF Health Africa (ESRC P1490/2023) and the University of Toronto (RIS Human Protocol Number: 46631).

### HEKA Research Initiative collaborative process

Figure 2 details the systematic process we are undertaking since establishing the research initiative. At a national level, all sexual and reproductive health community-based organizations would meet quarterly (peer assessment meeting) to foster peer-to-peer support across programs and enhance program delivery, through sharing best practices and local advocacy. During one such meeting in October 2019, the CBOs initiated conversations between our respective representatives. This happened in the context of long-standing academic collaborations where we felt excluded from some of the analysis-focused research components, and limited participation in key population mathematical modeling working groups. We (CBOs) set up monthly meetings thereafter, where we gathered information on the programmatic priorities of our organizations and assessed the baseline research expertise in our respective programs. We recognized the need for sustainable and rigorous capacity building within our research initiative, and as such, approached an academic colleague (SM) to co-design a skill development plan. We decided to organize an in-person meeting with all team members from respective CBOs to initiate skill development and set up a regular meeting agenda.

**Figure 2.**
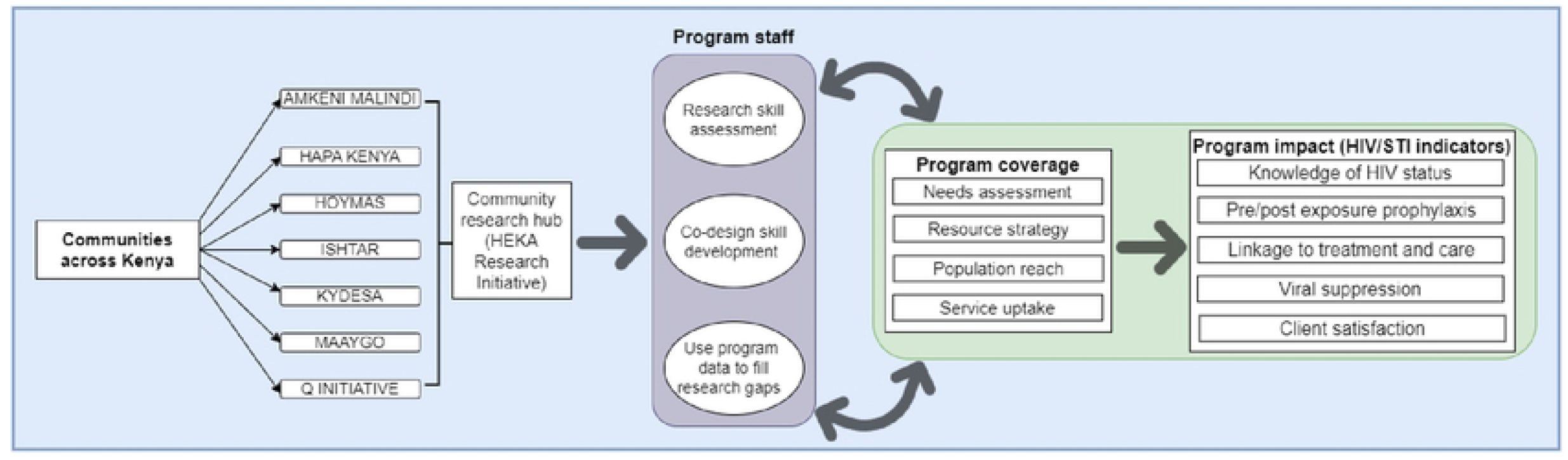
Systematic illustration of the HEKA Research Initiative collaborative process.

Organizing in-person meetings with representatives from across the country required financial resources, so we prioritized applying for funding. The COVID-19 pandemic brought our efforts to a pause, though it provided us to reassess our in-house tools in program delivery. We had generated routine or day-to-day person-level data through all the years of client contact. As such, we pursued funding to align our goal of holding regular in-person and virtual program staff research skill development with the use of routine programmatic data to guide our response along the HIV/STI prevention and treatment cascade. In 2023, we submitted the proposal, “Characterizing the HIV epidemic, prevention gaps, and opportunities among men who have sex with men in selected counties in Kenya using routinely collected program data before, during, and after the COVID-19 pandemic: a community-based research initiative” to Amref Health Africa and NACOSTI (National Commission for Science, Technology, and Innovation). The proposal was successful, and we obtained pilot funding to support our regular meetings and initiate the implementation of the proposed project.

At the center of the HEKA Research Initiative is that we provide optimum services and improve health outcomes for the communities we serve. Through quantitative skill development, we aim to use routine programmatic data to assess the needs of our communities (e.g., quantifying the client population longitudinally); map program initiatives and resources to population size (resource strategy); and use analytic tools to assess disease drivers and contextual factors influencing population reach to improve uptake of services. The HEKA Research Initiative systematic process is intended to be iterative.

### Collaborative meeting and outcomes

With the pilot funding, we organized the first official in-person meeting in November 2023. Beyond using this to set up our overall goal as a research group, we designed this as a three-day intensive workshop. This included interactive teaching and hands-on exercises on research concepts and methodology of quantitative analysis. The choice of three days and a mixture of didactic information sessions, hands-on learning, and small group projects was based on prior collaborative work and the Coordinating with Communities guidelines for improving community involvement in HIV prevention (agenda included in the Supplemental file)(29). For the workshop, we co-designed a skill development plan with our academic partners (NT, HM, and SM); this included an introductory session on the use of R Programming Language, an intuitive and open-access tool that will be helpful to achieve our analytic goals. The didactic component included sessions on study designs, formulating a “good” research question, types of variables, and principles for generating research study data from routine program data. The hands-on component involved preparing data from Microsoft Excel files as wide and long data formats and learning to use R to import data, using a dummy dataset. The team was split into three small working groups where we formulated three research questions, and each small team presented their work to the whole group, before questions were refined and specific objectives included.

At the end of our three-day workshop, we had drafted the overarching research goals for HEKA, identified three pertinent research questions (see Table 2), and set up a regular online meeting schedule for once a month with continuous communication via a new WhatsApp group.

**Table 2.** Preliminary HEKA research focus.

| **Sub-group research questions** |  |  |
| --- | --- | --- |
| **Program priority topic** | **Program-relevant research question** | **Counties** |
| Sexually transmitted infections (STIs) and condom supply | How did condom stockouts impact rates of sexually transmitted infections (STIs) among MSM who receive services at the CBOs? | Nairobi |
| oral HIV pre-exposure prophylaxis (PrEP) | What are the factors affecting the initiation, re-initiation, and retention of HIV oral PrEP among MSM between 18-24 years old and 35+ years old within the coastal region? | Kilifi, Mombasa, Trans Nzoia, and Uasin Gishu |
| Mental health among MSM living with HIV | What are the effects of mental health on viral suppression amongst MSM living with HIV in Kisumu and Nakuru, Kenya? | Kisumu and Nakuru |
| **Overarching HEKA research goals** |  |  |
| 1. Advance our scientific literacy in epidemiological research<br>2. Generate research questions and develop study designs<br>3. Learn the basics of an open-access software, R, to advance our data analysis & data visualization capacity<br>4. Develop a harmonized longitudinal study database using programmatic data<br>5. Develop a community of epidemiological research practice<br>6. Begin building expertise in community-based data science and community-based participatory mathematical modeling<br>7. Develop a research team entirely made up of and led by MSM living in Kenya |  |  |
| **Notes.** MSM: Men who have sex with men, CBO: Community-based organization, HEKA: <u>H</u>ealth <u>R</u>esearch <u>I</u>ntervention <u>K</u>uthamini <u>A</u>fya <u>Y</u>etu |  |  |

## Discussion

Since the initiation of our collective research initiative, we have informally engaged in reflective conversation to gather thoughts on what lessons we have learned so far in the process and based on challenges that have emerged, what our recommendations would be. After our first in-person meeting, we formalized the different thoughts we had in writing; the draft was reviewed by team members before final inclusion in this paper.

### Reflection on lessons learned

#### 1. Co-leadership framework

As community researchers, we often are token members of research teams, and so, would lead within the confines of the research plan of our academic colleagues. The HEKA Research Initiative has been uniquely successful because we have adopted a co-leadership framework where the direction of our work evolves through shared and delegated leadership across all seven community organizations. This way, we all feel equally engaged and create space for open conversation and consensus building around the direction to take in achieving our research goals. “*We are moving from data generators to also being data users*.*” “Let’s have a personal stake in this; I will renew my commitment – when we are doing our virtual engagement, this is “our baby” as a team*.”

#### 2. Accompaniment approach to learning about quantitative methods and R programming

We learned about the fundamentals of quantitative methods in research, securing us with a link between our intuition and observations and the standard terminologies that researchers use. This provides us with the tools to engage on a more level playing field when it comes to co-designing not only the research questions but also the analytic plans. The hands-on experience with data cleaning, data checks, and data management in R using an accompaniment approach meant that we were jointly learning, processing, and negotiating skills at every step. Often the needs of program decisions and research are faster than the learning process, but our team balanced this by pausing at each step to work out the data issues together. R is a language, like the language of communities, and walking side by side with communities and academics is how we learned to communicate with each other. “*Learning R has been a journey. It has been both physical workshops backed up by constant weekly virtual meetings both as a whole team as well as one-on-one meetings. This has led the team to continue being united, and hence the constant meetings have led to the process feeling like on-job training*.”

#### 3. Program data and rethinking HIV/STI indicators

Through direct interrogation of the data generated through programs, we are growing our ability to understand the data and begin to think of ways to identify gaps, both research-wise and also in the quality of the tools we currently use, in capturing relevant data, so as to make informed recommendations. “*Through this process, community-based organizations are not only producers of data to be consumed by researchers, but as we gain more skills, the researchers (coming from these KP organizations) can be able to analyze and interpret the data from program implementation and identify existing gaps in the program*.” “*This is a real capacity-building work – improving us from where we were. I can say I am not in the same place I was when we started doing this – especially with R. If we can continue the virtuals, this will continue to help my growth. This means we can analyze our own data much easier and know and see the gaps – not the other way around, where donors tell us what our gaps are and what to implement*.”

#### 4. Peer-to-peer mentorship and trust-building

The research initiative has been structured around multi-organization small working groups, where community-based organizations are grouped around common pertinent research questions. It was inherently built-in to support each other to meet the deliverables to advance our collective goal, such as co-learning and practicing R programming between sessions. This has fostered more peer-to-peer check-ins and other collaborative support between our programs, thus aiding in building trust, which is essential to our programs. “*I feel like everything is an opportunity for learning within HEKA. Want to give my thanks to HEKA leaders who made sure we can continue to have these experiences. This is also helping my organization improve, and I have been learning from other organizations and putting into practice what other organizations are doing*.”

### Challenges and recommendations for facilitating integrated community-led HIV/STI research

#### 1. Sustained engagement through funding

Community researchers also have a responsibility to run community-based programs and as such, go beyond to lead research in addition to work schedules. The HEKA Research Initiative has been a success because we have prioritized the compensation of staff to cover the time spent, including logistics for in-person meetings through small competitive grants from national agencies like Amref. HEKA’s plan around resource mobilization is to ensure these virtual meetings and in-person meetings are sustained all through the research period and beyond since there is limited funding and timeframe for the research.

#### 2. Difference in learning pace according to community researcher engagement level

Workshops are very few and limited to a few days; hence bringing the team together can be difficult with members coming from different professional backgrounds with different exposures (roles differ, for example, monitoring and evaluation staff and data clerks have more contact with data) to different tools. As a result, there is a difference in the learning pace. Therefore, we recommend allocating more days for these workshops as it would help ensure that all the team members can confidently participate in the overall process

#### 3. Socio-political climate

Due to the rising, state-sanctioned anti-LGBTQI+ movements, the in-person meetings were at many times disrupted or postponed to ensure the safety of our team. We leveraged virtual spaces such as WhatsApp communications and Zoom sessions, though these were sometimes interrupted during days when safety was a concern and staff could not make it to work. This was especially true for the organizations in regions most affected by anti-LGBTQI+ protests. These protests also delayed services and data collection and management.

#### 4. Long hours of travel

Long hours of road travel during the physical meetings contributed to some of the members being fatigued and hence less productive with the limited workshop days. With adequate funding, we would recommend booking flights for the in-person collaborative meetings, especially for more remotely-located team members.

#### 5. Data abstraction processes

Different times in data capturing using Excel workbooks have limited community researchers in data extracting/mining, which led to more time dedicated to data cleaning and harmonization. Due to limited exposure to the analytical tools, time was spent learning the tools and additional virtual meetings were required to troubleshoot and complete data abstraction and anonymization. We recommend building in more workshop and mentorship hours.

## Conclusion

This paper sets out to provide a novel example of community-led research using programmatic data, with insights into steps taken, reflections, and practical perspectives on sustainable and successful community-led collaborative research. We hope to inspire other community-based organizations in the HIV/STI research field to see that it is possible for funding organizations to facilitate the autonomy of community organizations to independently seek competitive grants towards the ownership and utilization of their programmatic data to inform service delivery.

We acknowledge the challenges involved in developing a community-led research platform with coordination across multiple geographically dispersed community organizations. The challenges we presented are not unique to our research initiative. Nonetheless, community-led collaborative research is a promising approach to mobilizing and addressing HIV/STI issues that are pertinent to our communities. HEKA boasts of multiple contributions to community-based research in Kenya:

1. It is the first time our community-based organizations have come together to interrogate quantitative data collected by our organizations to guide our response along the HIV/STI prevention and treatment cascade.
2. This is the first time our community organizations have in-depth and hands-on training on quantitative data analytic concepts and methods. Through co-designed skill development for our program staff, we are better equipped to actualize program science in HIV/STI coverage through the iterative use of routine program data to monitor and tailor program delivery and adapt to the evolving nature of the ecology of community programs in Kenya (e.g., a changing socio-political environment that shapes how we interact with service users and funders).

With our collaborative work underway (research agenda developed), some of our next steps include completing a scientific paper detailing the process we undertook in developing a harmonized database from the program data, which will facilitate rapid programmatic, community-led monitoring activities. HEKA demonstrates the potential for community-based and led research in the HIV/STI field. The model we present can serve as a blueprint for other community-based organizations aiming to lead collaborative or independent research and build capacity. As future steps, we will conduct analyses to address the research questions: root-cause quantitative analyses of mediators of new HIV and STI infections using a causal lens generated through lived experience; exploration of patterns and predictors of initiation, persistence, and re-initiation of HIV pre-exposure prophylaxis across diverse geographic regions within Kenya; exploring the impact of disruption in the supply of commodities on STI diagnoses, condoms, and lubricants; and the development of a group governance document (for sustainability). HEKA will also begin building fundamental understanding of mathematical modeling, as we plan to implement community-based participatory mathematical modeling.

## Data Availability

Not applicable

## Acknowledgment

We thank each of our respective community-based organizations for supporting the HEKA initiative. We also thank the Kenya Key Population Consortium for encouraging the HEKA initiative. NT is supported by the St. Michael’s Foundation Angel’s Den Doctoral Scholarship. SM is supported by a Tier 2 Canada Research Chair in Mathematical Modeling and Program Science. RL is supported by a Tier 2 Canada Research Chair in Global Intervention Politics and Social Transformation.

## Box 1.

List of services offered by the community-based organizations

- HIV testing services
- HIV care and treatment
- Tuberculosis screening and referral
- Risk reduction counseling
- Provision of pre-and post-exposure prophylaxes
- Condom/lubricant education and distribution
- One-on-one group peer outreach HIV and sexual and reproductive health education and counseling.
- Referral for harm reduction services for key populations using drugs
- Support and referrals for paralegal services
- Sexually transmitted infection screening and treatment
- Psychosocial support groups
- Nutritional assessment and referral
- Mental health assessment and referral
- Gender-based violence assessment and referral
- In-house social activities to increase retention in care and treatment
- Crisis response
- Security and safety literacy
- Economnic empowerrnent, financial literacy and skill-building
- Policy advocacy

